# Metabolic modulation of brain network dynamics during alcohol withdrawal

**DOI:** 10.64898/2026.10.07.26364946

**Authors:** Julia K. Brynildsen, Ehsan Shokri-Kojori, Corinde E. Wiers, Nora D. Volkow, Dani S. Bassett

## Abstract

Chronic heavy alcohol use shifts brain metabolism from glucose toward acetate utilization. Sudden decreases in acetate following alcohol cessation in individuals with alcohol use disorder (AUD) may create an energetic imbalance that contributes to craving and withdrawal. A recent randomized study found that, compared with a standard American (SA) diet, a ketogenic diet (KD) during alcohol detoxification reduced withdrawal severity and craving and increased ketones and glutamate in the dorsal anterior cingulate cortex. These findings suggest that ketones may support metabolic adaptation during alcohol withdrawal, but the effects of altered brain metabolism on neural dynamics and alcohol cue reactivity remain unknown. Here, we used a control theoretic approach to test whether shifts in cerebral energy substrate utilization influence large-scale network dynamics through altered information flow. We analyzed fMRI cue-reactivity scans collected after one, two, and three weeks of inpatient treatment during presentation of alcohol, food, and neutral cues. Whole-brain effective connectivity was estimated using Shannon transfer entropy applied to whole brain atlas-aligned fMRI time series. Assuming neural activity flow along effective connections, we identified distinct shifts in the state-transition landscape in participants receiving the KD compared with the SA diet. Increases in serum *β*-hydroxybutyrate (BHB) were associated with changes in state-transition landscape organization, which were in turn associated with reduced cue-induced alcohol craving. These findings link metabolic adaptation to large-scale neural dynamics and clinically relevant behavior and suggest that cerebral energy substrate availability may shape behavior by altering the organization of large-scale neural dynamics.

## Introduction

Alcohol use disorder (AUD) is a chronic, relapsing condition characterized by the inability to control or stop alcohol use despite negative social, occupational, or health consequences [1, 2]. Chronic alcohol consumption in AUD is thought to induce a persistent bioenergetic shift in the brain, whereby glucose metabolism decreases, and metabolism of acetate, an alcohol metabolite, increases [3–5]. During alcohol withdrawal, decreased acetate availability may induce an “energy-depleted” state in the brain that contributes to heightened craving and symptoms of withdrawal. Dysregulated brain metabolism is evidenced by altered concentrations of glutamate and glutamine [6], key components of the glutamate/GABA-glutamine cycle that contribute to the maintenance of neurotransmitter homeostasis [7, 8]. Disrupted neurotransmitter homeostasis across multiple systems in alcohol withdrawal is associated with neuronal hyperexcitability [9], which alters communication between brain regions. A clear understanding of how brain metabolism may influence large-scale brain network dynamics to modulate alcohol craving is key for the development of novel therapeutic approaches to mitigate relapse.

Ketogenic diets provide an alternative metabolic substrate in the form of ketone bodies, including *β*-hydroxybutyrate (BHB), which can be utilized by the brain during periods of reduced glucose availability [10]. Thus, ketogenic dietary intervention provides a means of altering systemic metabolism during alcohol withdrawal, while circulating BHB provides a measurable index of the resulting metabolic response. Importantly, this metabolic perturbation via dietary intervention in AUD provides an opportunity to study the link between peripheral metabolic state and large-scale brain function. Understanding whether individual differences in BHB response are associated with changes in brain network organization may therefore provide insight into how metabolic interventions influence neural dynamics and behavior.

Traditional approaches to understanding the neural circuitry underlying the craving of alcohol and other substances have focused on correlations between regional activation (e.g., within parts of the brain’s reward system) and behavioral measures (e.g., craving) in response to cues [11–14]. However, it is increasingly recognized that AUD and other substance use disorders are maintained by large-scale changes in brain network connectivity and communication [15]. In cue reactivity, ensembles of brain networks function in coordination to engage attention, salience, cognitive control, and reward, among other functions [16]. In recent years, the brain’s salience network has emerged as a key modulator of reactivity to alcohol cues, situated at the intersection of large-scale networks associated with AUD [17]. The dorsal anterior cingulate cortex (dACC), a key node of the salience network, influences dopamine release within the ventral striatum (VS), a reward-encoding region, in response to alcohol cues [18]. In healthy individuals, dopamine release in the VS is inversely correlated with metabolic activity in the anterior cingulate cortex (ACC), but this relationship is disrupted in alcohol dependence [19].

Network control theory (NCT) provides a means of deriving mechanistic insights into how whole-brain connectivity gives rise to functional brain states associated with craving. In the context of NCT, control energy is defined as the cognitive effort associated with driving the brain to a particular state, and controllability is operationalized as a region’s capacity to enact changes in whole-brain state [20]. Recent evidence indicates a strong relationship between regional glucose metabolism and control energy required to transition between states defined by activity within networks of functionally-associated brain regions [21]. Thus, a control theoretic approach could be useful for predicting the influence of metabolic shifts in the brain on cognition and behavior.

In this study, we seek to link diet-induced metabolic influences on brain dynamics associated with cue reactivity by leveraging a unique multimodal dataset from a population of treatment-seeking patients with AUD (n=33) [5]. Participants in the study were inpatients in a detoxification unit and randomized to either receive a ketogenic (KD) or standard American (SA) diet for three weeks following alcohol discontinuation. Ketogenic diets, which regulate brain metabolism during withdrawal by providing alternative energy substrates, have shown promise as a strategy for downregulating craving and other symptoms of withdrawal, and offer a means of experimentally manipulating metabolic state to test the proposed control-theoretic relationships [4, 5, 22]. During each week of the study period, participants underwent an fMRI scan during which they reacted to alcohol cues and a magnetic resonance spectroscopy scan during which concentrations of metabolites were measured in the dACC (Figure 1). Relative to the SA group, the KD group showed increased levels of glutamate and glutamine in the dACC, increased dACC reactivity, and decreased craving in response to alcohol cues. Participants in the KD group also needed significantly fewer benzodiazepines during the first week of withdrawal than participants in the SA group [5]. Additionally, analysis of fMRI cue-reactivity data from the same participants showed lower expression of a neurobiological craving signature in the KD compared with the SA group in response to alcohol relative to food cues [23].

**Figure 1:**
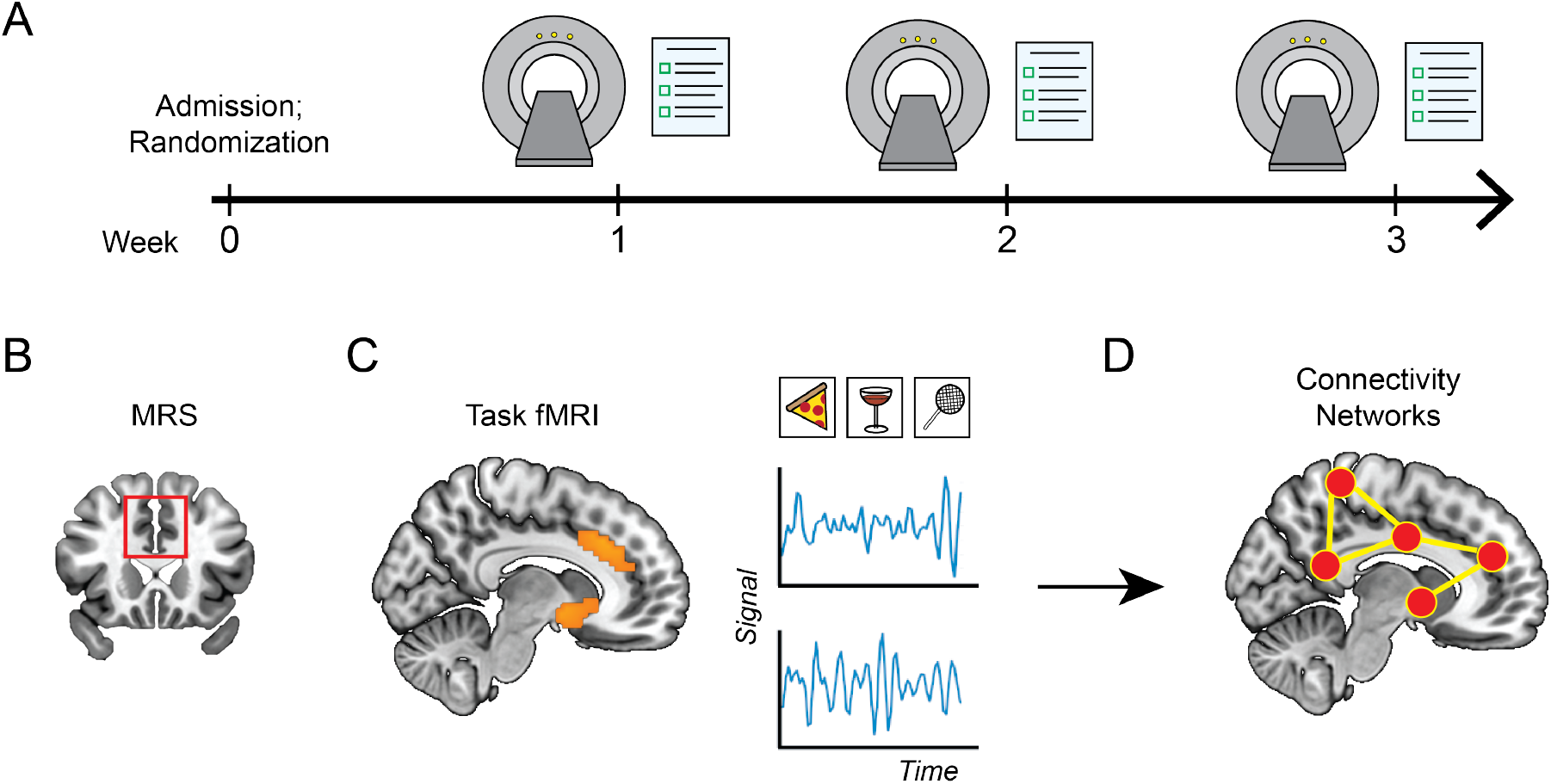
Study overview. **(A)** Participants were assigned to ketogenic diet (KD) or standard American diet (SA) groups prior to undergoing three weeks of fMRI and magnetic resonance spectroscopy (MRS) scans. **(B)** Glutamate and glutamine were measured via MRS in the dACC. **(C)** fMRI data were acquired while participants reacted to food, alcohol, or neutral cues in the scanner. **(D)** Shannon transfer entropy was used to derive effective connectivity networks from fMRI time series data at each time point.

Here, we used information theory and NCT to examine relationships among metabolic state, large-scale brain dynamics, and alcohol cue reactivity during withdrawal. We addressed four related questions. First, we tested whether metabolic modulation through dietary intervention was associated with changes in network-level information flow in the brain. Second, we examined whether diet condition altered the organization of the energetic landscape governing transitions between large-scale brain states. Third, we tested whether changes in this organization were associated with changes in alcohol cue reactivity, focusing on alcohol wanting. Finally, we examined whether changes in circulating BHB were associated with changes in the organization of the transition-energy landscape, and whether changes in this organization statistically accounted for the relationship between BHB and alcohol wanting. This study links changes in metabolic state with changes in alcohol cue reactivity during withdrawal, providing a framework for predicting the impact of metabolic perturbations on recovery.

## Methods

### Participants

Data were previously acquired as part of a randomized clinical study examining the effects of a ketogenic diet on alcohol withdrawal and alcohol-related outcomes in individuals with AUD [5]. The original study enrolled 53 treatment-seeking inpatients at the National Institute on Alcohol Abuse and Alcoholism (NIAAA) to undergo detoxification. Of these, 33 participants completed the 3-week dietary intervention (19 assigned to the ketogenic diet and 14 to a standard American diet); 20 participants withdrew before completing the intervention. The original study was approved by the Institutional Review Board at the National Institutes of Health, and all participants provided written informed consent. Detailed information regarding participant recruitment, screening, eligibility criteria, clinical characteristics, and the study protocol has been reported previously [5].

### Dietary intervention

Within two days of admission, participants were randomized into ketogenic diet (KD) (n=19) or standard American diet (SA) (n=14) regimens. The KD diet comprised 80% calories from fat, 15% calories from protein, and 5% calories from carbohydrates; the SA diet consisted of 35% calories from fat, 15% calories from protein, and 50% calories from carbohydrates. Each diet consisted of a shake and a solid ketogenic snack at each meal, and participants were blinded to the diet condition. As confirmation of KD effects, blood BHB and urinary acetoacetate (AcAc) were measured daily, with participants in the KD group showing significantly higher levels of both ketone metabolites relative to SA participants [5].

### Neuroimaging data

Structural and functional MRI data were acquired using a 3.0-T Siemens Magnetom Prisma scanner (Siemens Medical Solutions USA Inc., Malvern, PA) with a 32-channel head coil, as previously described [5]. fMRI cue reactivity scans were acquired using a standard echo-planar imaging sequence (repetition time = 1.5 s, echo time = 30 ms, flip angle = 70^*°*^, 36 slices, 3 *×* 3 *×* 4 mm voxels) during weeks 1, 2, and 3 of the study. During each cue reactivity scan, a total of 40 alcohol, 40 appetitive food, and 40 neutral cues were presented over the course of three runs in an event-related design. Each cue was presented for 750 ms, and the total task duration, including all runs, was approximately 13.5 min [5, 24]. Following each MRI session, participants rated the cues for subjective wanting in response to the question “How much do you want to consume this right now?” using a 7-point Likert scale ranging from 0 (not at all) to 6 (extremely) [5]. In the present analyses, alcohol cue reactivity was quantified as mean wanting for alcohol cues relative to neutral cues (alcohol *−* neutral wanting).

For structural and functional data preprocessing, the minimal preprocessing pipeline developed for the Human Connectome Project (HCP) was used [25]. Briefly, the HCP functional preprocessing includes correction of spatial distortions, correction for subject motion, registration of functional images to the individual structural image, intensity normalization, and alignment to the standard MNI space. The minimal HCP pipeline is designed to preserve the native temporal information in the fMRI data and does not, by itself, include additional temporal filtering, nuisance regression, or motion censoring [25]. Preservation of temporal structure was particularly important for the present analyses because the regional BOLD time series were subsequently used to estimate directed interregional dependencies using transfer entropy. Transfer entropy depends explicitly on the temporal ordering and lagged relationships among signals, such that alterations to their temporal structure may influence estimates of directed information transfer [26].

Regional fMRI time series were extracted using the ‘3dNetCorr’ function in AFNI [27] from the 200-region Schaefer cortical atlas [28] and 50-region Tian subcortical atlas [29], yielding 250 cortical and subcortical regions in total. For each region, the mean BOLD signal across voxels within the corresponding parcel was extracted at each time point to generate regional time series.

### Derivation of whole-brain effective connectivity

Effective connectivity (EC) provides a means of statistically inferring activity flow across brain regions [30]. In the context of this study, we calculated Shannon transfer entropy (TE) to estimate directed information flow between regional fMRI time series. TE is an information-theoretic approach that quantifies the extent to which the past of one time series improves prediction of the future of another time series beyond the information contained in the target time series’ own past [26]. For two regional BOLD time series *X* and *Y*, TE from *Y* to *X* can be expressed as

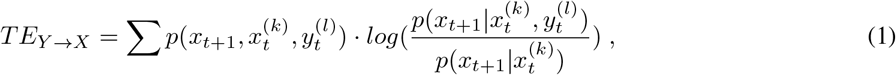

where 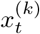 and 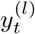 denote the histories of *X* and *Y* over *k* and *l* previous time points, respectively; *p* denotes probability; and the vertical bar denotes conditional probability. In the present analysis, we used the default first-order Markov history implemented by the package `RTransferEntropy` [31] (*k* = *l* = 1). The BOLD time series were entered directly into the *calc_te()* function and were discretized internally using the package’s default empirical-quantile procedure before estimation of Shannon TE.

For each participant, scan session, and run, TE was estimated for every ordered pair of the 250 brain regions defined by the combined Schaefer 200-region cortical and Tian 50-region subcortical parcellations. The resulting 250 x 250 matrix represented directed information flow, with target regions indexed by rows and source regions indexed by columns; thus, *A*_*ij*_ represents information flow from region *j* to region *i*. Because the primary goal was to characterize changes in the organization of directed information flow across the three study time points, TE matrices were estimated separately for each participant at each week of the intervention and averaged across the three runs. We computed the Spearman correlation of directed edge weights between every pair of time points for each participant to obtain a measure of edge weight stability.

### Network control theory analyses

Network control theory (NCT) is a systems engineering approach that determines which perturbations can drive a system to a desired state [32, 33]. Here, we used NCT to compute the energetic cost associated with transitioning between activation of distinct functional subnetworks. Specifically, we modeled neural dynamics as a linear, continuous-time, and time-invariant system:

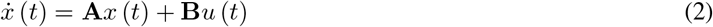

where *x* (*t*) is the vector of activity states across the *N* brain regions, **A** is the *N × N* directed network adjacency matrix, *u* (*t*) is the external control input, and **B** specifies the brain regions receiving control input in order to drive the state change of the network. In the present analyses, control input was available to all brain regions, such that **B** is the *N × N* identity matrix.

For each participant, time point (1, 2, and 3 weeks), and study run (A, B, and C), the directed transfer entropy matrix was used as the dynamics matrix **A**. Prior to computing the optimal control energy, **A** was normalized to promote numerical stability [34]. We then estimated the minimum control input required to transition between predefined brain states over a fixed time horizon of *T* = 3. Control energy was calculated from the squared magnitude of the optimized control input over the simulated trajectory.

Brain states were defined by activity localized to each of the 17 Yeo-Krienen functional subnetworks [35] or subcortical regions. Thus, for each participant, we computed the energetic cost associated with every possible pairwise transition among the functional subnetworks. For example, Fig. 3A represents the transition from activation of the somatomotor network 1 to activation of visual network 2. The initial state *x* (0) is a 250 *×* 1 vector containing ones in regions of somatomotor network 1 and zeros elsewhere; the final state *x* (*f*) is a 250 *×* 1 vector containing ones in regions of visual network 2 and zeros elsewhere.

Recent work has demonstrated the validity of linear models for accurately capturing neural dynamics at the macroscale [36] and has established the similarity of the control theoretic framework to communication models used to explain the transfer of signals between neurons [37]. Together, these studies support the use of linear network control theory as a biologically plausible framework for quantifying the energy cost associated with transitions between large-scale functional brain states.

### Dimensionality reduction of state transition matrices

To assess changes in the structure of the state transition landscape across time, we performed a singular value decomposition (SVD) of each participant’s state transition matrix at each time point. Briefly, SVD factorizes a matrix into three elements; left-singular vectors, singular values, and right-singular vectors, with the singular values quantifying the contribution of each corresponding component to the overall structure of the matrix. We focused on the variance explained by the first singular value (VE1), which captures the dominant organizational component of the state transition matrix. Thus, higher VE1 values indicate that a greater proportion of the structure of the transition landscape is captured by a common underlying pattern, whereas lower values indicate a more distributed organization across multiple patterns. Prior to performing SVD, the control energy values associated with each state transition were *z*-scored across time points for each participant.

### Statistics

To examine the effects of diet condition and time interval on TE network stability, we fit a generalized least squares model with fixed effects of diet condition, time interval (weeks 1–2, weeks 2–3, or weeks 1–3), and their interaction. To account for the nonindependence of the three within-participant stability estimates, including overlap among intervals, we specified an unstructured within-participant correlation matrix. In the final model, residual variance was also allowed to differ across intervals. To examine the effects of time and diet condition on VE1, we employed a linear mixed effects model that included fixed effects of diet condition, week, and their interaction, and participant ID as a random effect. Spearman’s correlations were used to assess associations between changes in the VE1 and changes in alcohol wanting, as well as associations between changes in BHB, VE1, and alcohol wanting from weeks 1 to 3.

To examine whether changes in VE1 statistically accounted for the association between changes in BHB and changes in alcohol wanting, we conducted an indirect-effects analysis using structural equation modeling (SEM) with the R package lavaan [38]. The model specified change in VE1 as a function of change in BHB and change in alcohol wanting as a function of change in both VE1 and BHB. The indirect effect was defined as the product of the BHB-to-VE1 and VE1-to-wanting path coefficients. Standard errors and 95% confidence intervals for model parameters were estimated using 5,000 nonparametric bootstrap samples. A *p <* 0.05 was considered significant across all analyses.

## Results

### Metabolic influence on the topology of information flow in the brain

We first set out to test the hypothesis that metabolic modulation via a dietary intervention during alcohol withdrawal alters network-level information flow in the brain. Transfer entropy (TE) is an information-theoretic approach that can be used to quantify the amount and direction of information flow between brain regions [39–41]. We derived transfer entropy from fMRI time series at each time point to construct effective connectivity networks in which nodes represent brain regions and edges represent information flow between them. To understand how changes in brain metabolism impact interregional communication, we focused on examining how information flow evolves across time in both treatment groups.

We began by examining the group-level stability of edge weights in transfer entropy networks. Figure 2A represents group-level edge stability, where denser edges are those exhibiting greater temporal preservation of their relative TE strength across weeks. A generalized least squares model accounting for within-participant dependence among the overlapping interval estimates revealed a significant main effect of diet condition on TE network stability, with lower stability in the KD group than the SA group (*F* (1, 87) = 6.095, *p* = 0.016). There was no significant main effect of interval (*F* (2, 87) = 3.066, *p* = 0.052) and no diet condition *×* interval interaction (*F* (2, 87) = 0.370, *p* = 0.692) (Fig. 2B). Thus, directed network topology underwent greater reconfiguration across withdrawal in the KD group, whereas the SA group retained a more stable pattern of directed information flow across weeks. Importantly, empirical TE networks exhibited substantially greater temporal stability than surrogate networks in which interregional temporal alignment was disrupted, supporting the presence of structured and reproducible directed information flow in the empirical data (Supplementary Fig. 1).

**Figure 2:**
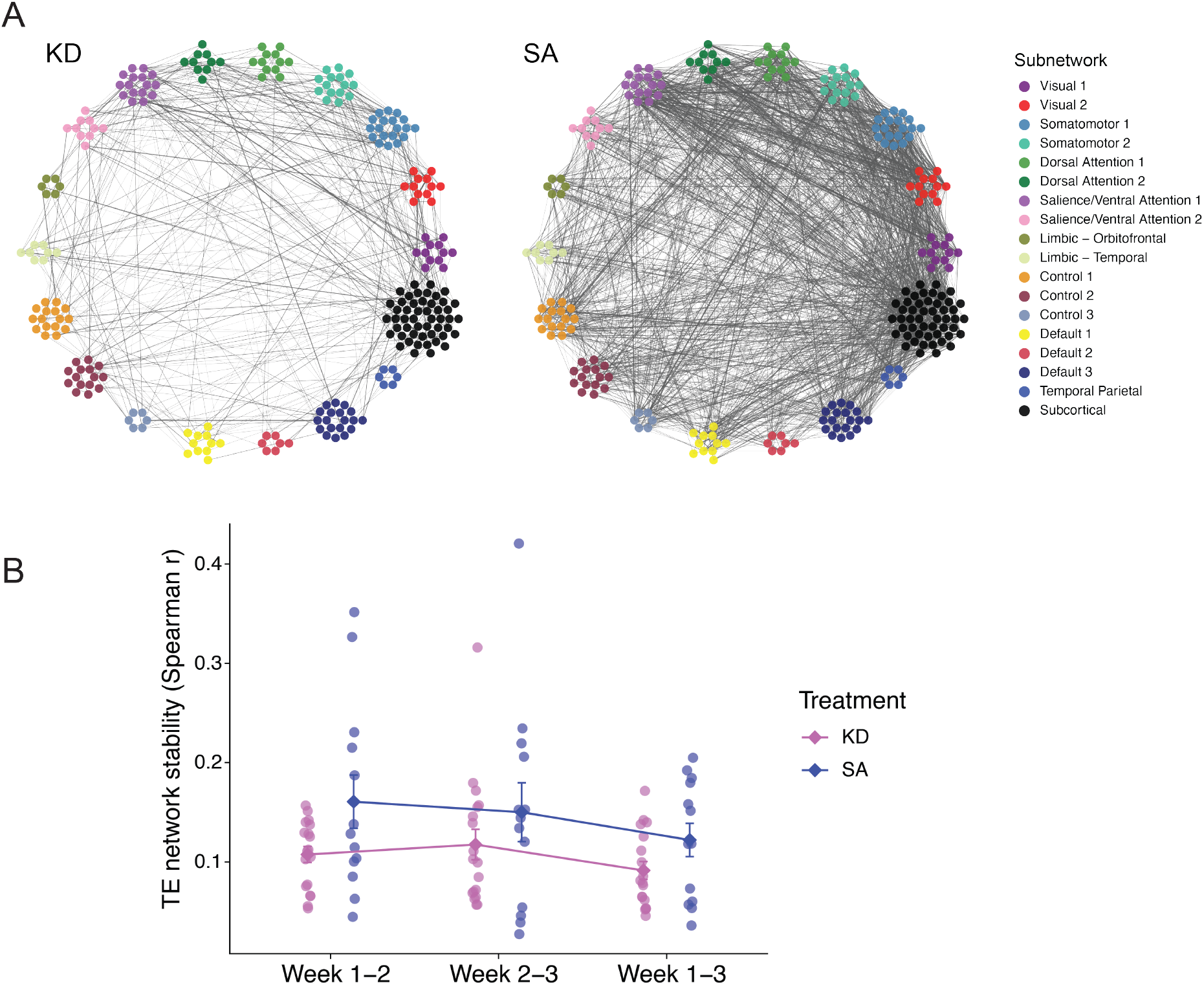
Temporal stability of directed information flow during alcohol withdrawal. **(A)** Network graphs representing the change in information flow between each pair of brain regions over the course of the study. Edges depict connections exhibiting the greatest temporal preservation of their relative TE strength across weeks. For visualization, the same stability threshold, defined as the 97.5th percentile of pooled KD and SA edge stability values, was applied to both groups. Nodes represent brain regions, which are clustered based on their identities in subnetworks defined a priori by Yeo-Krienen. **(B)** Stability of participant-level network topology. Stability reflects the Spearman correlation across edges for each participant between every pair of time points. Points represent individual participants, and error bars represent *±* SEM.

**Figure 3:**
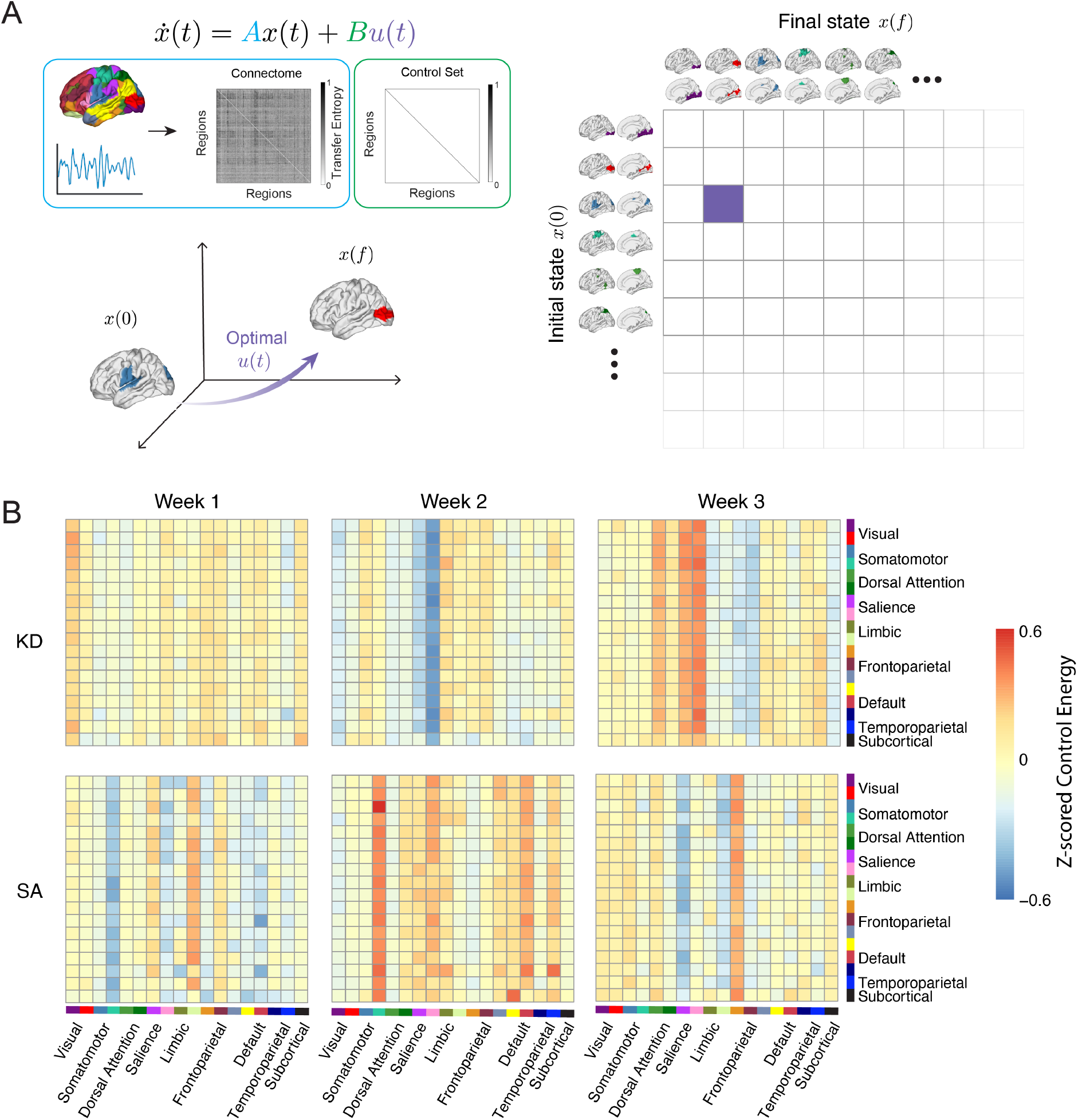
Energetic cost of transitions across large-scale brain networks. **(A)** Using a linear, continuous time, and time-invariant model to approximate neural dynamics, we computed the control energy required to transition between all possible pairs of brain states. Each participant’s 250 *×* 250 connectome **A** represents effective connectivity (transfer entropy), and control inputs were given to all brain regions. Each initial state *x*(0) and final state *x*(*f*) were defined as a 250 *×* 1 vector containing ones within a specific functional network and zeros elsewhere. **(B)** This approach was used to compute the log sum energy cost associated with transitioning between every possible pair of functional states in participants who received a KD (top row) or SA (bottom row) diet. Warmer colors indicate relatively higher control energy and cooler colors indicate relatively lower control energy. Control energy values were standardized (z-scored) across time within participant, such that colors reflect relative temporal changes in the energetic cost of each state transition.

### Energetic cost of transitioning among large-scale brain networks

We next sought to quantify the influence of diet-induced changes in interregional information flow on large-scale neural dynamics. We computed the log sum energy cost associated with transitioning between neural states that reflect activation of distinct functional brain networks, assuming activity flow along directed connections (Fig. 3A). Thus, we considered transitions from an initial state of activity in each functional network to activity in every other functional network. Each state vector contained ones in regions belonging to a given functional network and zeros elsewhere.

We generated heatmaps reflecting the energy cost associated with transitioning between every possible pair of functional networks (Fig. 3B). Higher values indicate transitions that require greater modeled control input relative to that participant’s other time points, whereas lower values indicate transitions that require less control input. Thus, the vertical and horizontal patterns visible in Fig. 3B reflect coordinated changes in the relative energetic cost of transitions into or from particular functional networks. For example, the relatively low values involving the salience network in the KD group at week 2 indicate a transient reduction in the modeled cost of those transitions, whereas the relatively high values involving somatomotor networks in the SA group at week 2 indicate an increase in their relative transition cost. Because control-energy values were standardized across time within participant, these patterns should be interpreted as temporal changes in transition cost rather than absolute differences in energetic demand between networks.

We performed singular value decomposition (SVD) [42] to assess the impacts of time, diet condition, and their interaction on the structure of the transition energy landscape. In essence, SVD is a mathematical approach that breaks down data matrices into constituent factors [43]. Here, we focus on the variance reflected in the first component (VE1), which captures the maximum possible variance. In the context of this study, VE1 can be thought of as a measure of the degree of structure in the overall transition landscape.

A linear mixed effects model was fitted to examine the effects of treatment group, time point, and their interaction on the linear trajectory of VE1, with a random intercept for participant. There was no significant main effect of group (*F* (1, 89.99) = 0.84, *p* = 0.361) or week (*F* (1, 61.18) = 1.99, *p* = 0.164). However, there was a significant time *×* treatment interaction (*F* (1, 61.18) = 5.20, *p* = 0.026), indicating that the change in VE1 over time differed between the two groups. Follow-up simple-slope analyses showed no significant change in VE1 over time in the KD group (*β* = 0.0033, 95% CI [*−*0.0065, 0.0132], *p* = 0.502), whereas VE1 significantly decreased over time in the SA group (*β* = *−*0.0142, 95% CI [ *−*0.0259, *−*0.0024], *p* = 0.019) (Fig. 4). Higher VE1 values indicate that transitions are increasingly governed by a common organizational pattern rather than many unrelated patterns, whereas lower values suggest a more fragmented state transition landscape.

**Figure 4:**
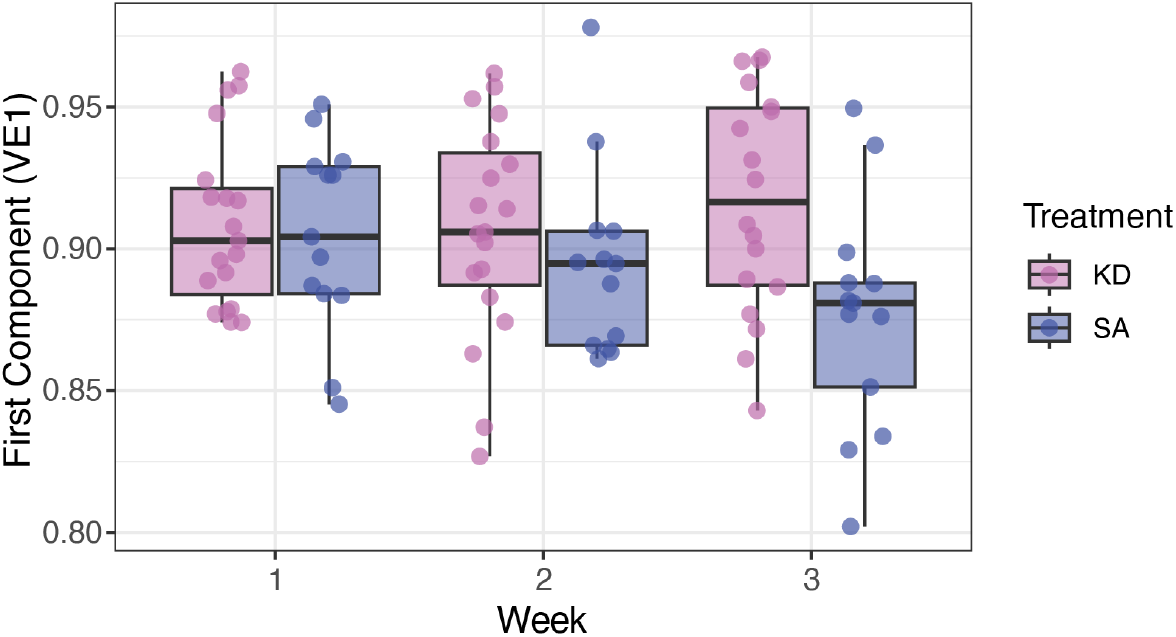
Dimensionality reduction of transition energy landscapes across time. Variance explained by the first component (VE1) following singular value decomposition of the state transition landscape for each participant at each time point. Points represent individual participants, and box plots show median, interquartile range, and 1.5 *×* IQR whiskers.

### Metabolic associations with brain network dynamics and alcohol wanting

To understand whether differences in brain network dynamics could be related to treatment outcomes, we examined the relationship between the change in VE1 and the change in alcohol wanting between weeks 1 and 3 of the study. A Spearman correlation identified a significant negative relationship between the change in VE1 from weeks 1 to 3 and the change in alcohol wanting (*ρ* = *−*0.54, *p* = 0.003) over the same time period (Fig. 5A). This suggests that a more structured state transition landscape may be associated with lower levels of alcohol craving during the cue reactivity task.

**Figure 5:**
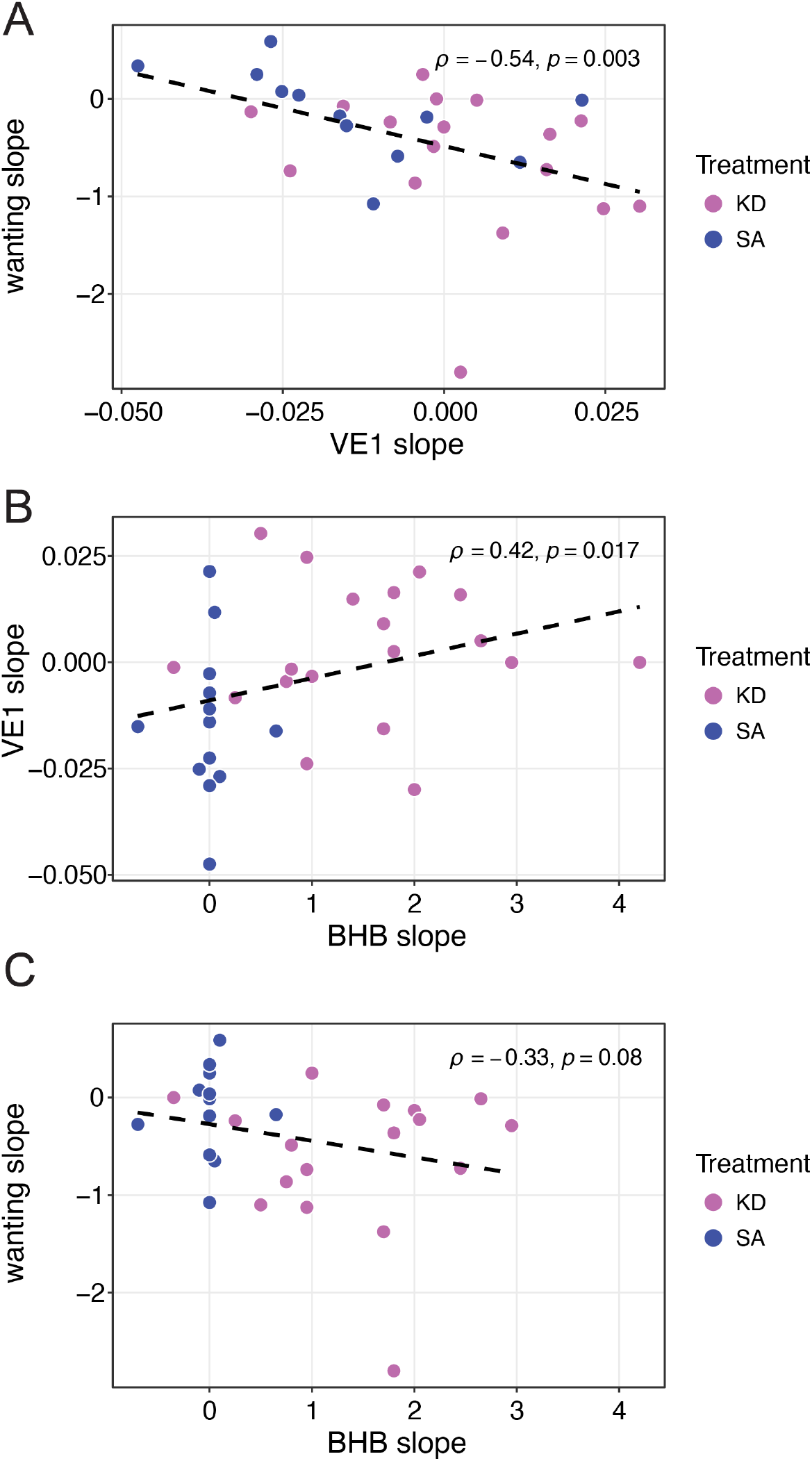
Relationships between BHB, VE1, and alcohol wanting. Spearman correlations between the change in VE1 and the change in alcohol wanting **(A)**, the change in BHB and the change in VE1 **(B)**, and the change in BHB and the change in alcohol wanting **(C)** during the three-week treatment period. Points represent individual participants.

Given the observed relationship between changes in VE1 and alcohol wanting, we next examined whether changes in circulating *β*-hydroxybutyrate (BHB) were associated with changes in VE1 and whether VE1 statistically accounted for the relationship between BHB and alcohol wanting. Across participants, changes in BHB were positively associated with changes in VE1 (Spearman’s *ρ* = 0.42, *p* = 0.017) (Fig. 5B). Changes in BHB also showed a negative, although nonsignificant, association with changes in alcohol wanting (*ρ* = *−*0.33, *p* = 0.080) (Fig. 5C).

To better understand the relationships among these variables, we tested an indirect-effects model in which change in BHB predicted change in VE1, which in turn predicted change in alcohol wanting. BHB change was positively associated with VE1 change (*β* = 0.333, *p* = 0.037), whereas VE1 change was negatively associated with alcohol wanting after accounting for BHB change (*β* = *−*0.402, *p* = 0.001). The direct association between BHB and wanting after accounting for VE1 was not significant (*β* = *−*0.122, *p* = 0.530). The estimated indirect effect was negative (standardized indirect effect = *−*0.134), with a bootstrap 95% confidence interval of [ *−*0.219, *−*0.004]. Together, these findings are consistent with an indirect association between increases in BHB and reductions in alcohol wanting through changes in VE1. Because the slopes of BHB, VE1, and alcohol wanting were each calculated over the same three-week time period, this analysis does not establish temporal or causal mediation.

## Discussion

In this study, we use information theory and network control theory to examine how diet-induced metabolic modulation during alcohol withdrawal is associated with changes in large-scale brain dynamics and alcohol cue reactivity. Specifically, we used transfer entropy to identify differences in the trajectory of directed information flow between participants receiving a ketogenic diet (KD) and those receiving a standard American diet (SA). These changes were accompanied by differences in the organization of the brain state transition landscape: although there was no overall difference in the structure of the transition landscape between groups, the trajectory of its organization over time differed between groups. Specifically, VE1 remained relatively stable over time in the KD group but decreased significantly in the SA group. Importantly, changes in this measure of transition landscape organization were associated with changes in alcohol wanting and statistically accounted for an indirect association between increases in BHB and reductions in wanting. Together, these findings suggest that metabolic modulation during alcohol withdrawal is associated with changes in the organization of large-scale neural dynamics, and that these changes are related to behavioral effects of alcohol withdrawal.

The group-level differences in transfer entropy networks afford insight into the effects of dietary intervention on interregional communication during withdrawal. Relative to the SA group, the KD group exhibited lower stability in edge weights over the course of treatment. Transfer entropy measures the extent to which the past of one regional time series improves prediction of another and therefore provides an estimate of directed statistical dependence between regions [26]. The present findings indicate that the dietary intervention is associated with greater reorganization of large-scale information flow during withdrawal, providing a potential network-level substrate through which altered brain metabolism could influence neural function.

Our network control analysis provides a means of characterizing the functional implications of these changes in information flow. Within the network control theory framework, brain states may be conceptualized as points along an energy landscape. The ease with which this landscape is traversed may be conceptualized as control energy– state transitions that are more difficult to complete require more control energy, whereas transitions that are easier to complete require less. Previous work has shown that the control energy landscape is malleable and may shift in response to external inputs, such as pharmacological modulation [44, 45], or in response to internal cognitive shifts, such as mindful attention [46] or habit learning [47]. Here, we demonstrate changes in the model-derived state-transition landscape associated with diet-induced metabolic perturbation during alcohol withdrawal.

By treating TE networks as the dynamics matrix of a linear control system, we estimated the relative cost of transitioning between states defined by activity in large-scale functional networks. We found that the structure of this landscape changed differently over time in the two dietary groups. Specifically, the variance captured by the first singular value (VE1) remained relatively stable over time in the KD group and decreased in the SA group, resulting in distinct group-specific trajectories. More broadly, large-scale brain activity is known to exhibit low-dimensional and hierarchical structure, such that a relatively small number of dominant modes or gradients can account for substantial variance in functional organization [48–50]. Viewed in this context, VE1 provides a measure of the extent to which the organization of the state-transition landscape is dominated by a common mode rather than distributed across multiple independent dimensions.

An increase in VE1 therefore indicates that a larger proportion of the structure of the transition landscape can be described by a common underlying pattern. In this sense, maintenance of VE1 in the KD group, relative to its decline in the SA group, is consistent with preservation of a more low-dimensional or coordinated organization of the relationships among the costs associated with transitions between large-scale brain states. This interpretation is also broadly consistent with evidence that cerebral energetic state can influence large-scale functional organization. Ketosis induced through diet or exogenous BHB has been shown to increase the stability of functional brain networks in healthy adults [51], and ketogenic interventions have been associated with increased functional connectivity in attention networks that covaries with regional ketone uptake [52]. More generally, regional glucose metabolism is systematically related to functional network topology and is particularly elevated in highly connected network hubs [53, 54]. The present findings extend this work by suggesting that metabolic modulation may influence not only the strength or stability of functional coupling, but also the organization of the energetic relationships governing transitions among large-scale brain states.

The relationship between changes in the transition landscape and alcohol cue reactivity provides further support for the functional relevance of this network-level organization. Changes in VE1 were negatively associated with changes in alcohol wanting, such that participants showing greater increases in the organization captured by the dominant component also showed greater reductions in wanting. Changes in BHB were also positively associated with VE1, and there was an indirect association between increases in BHB and reductions in wanting through changes in VE1. Cue reactivity requires coordinated engagement of multiple systems involved in perception, attention, salience, reward, and cognitive control [16]. A more structured transition landscape could therefore reflect a neural system in which transitions among large-scale functional states are governed by more consistent organizational constraints, potentially reducing the degree to which salient alcohol cues are associated with changes in the broader configuration of neural activity.

The metabolic context of these findings is particularly relevant during alcohol withdrawal. Chronic heavy alcohol exposure is associated with altered cerebral metabolism, such that acetate metabolism increases and glucose metabolism decreases [3, 4, 55]. During withdrawal, the abrupt reduction in acetate availability may therefore place additional metabolic demands on the brain at a time when neurotransmitter homeostasis and neuronal excitability are already disrupted. Withdrawal-associated hyperexcitability could, in turn, interfere with the coordinated activity of distributed neural systems, providing one potential link between altered cellular physiology and the large-scale network reorganization observed here. From this perspective, improved metabolic support during withdrawal could contribute to stabilization of neural activity and thereby alter the organization of information flow and state transitions across the brain. The parent clinical study found that the KD intervention was associated with higher ketone levels, increased dACC glutamate and glutamine, reduced alcohol craving, and reduced benzodiazepine requirements during the first week of withdrawal [5]. The present findings extend those observations by suggesting that the consequences of dietary metabolic modulation may be detectable at the level of large-scale brain network dynamics. Rather than influencing only regional activation patterns, provision of alternative energy substrates may alter the organization of information flow across distributed neural systems.

Ketone bodies provide a plausible metabolic substrate for such effects. Ketogenic diets may provide an efficient alternative fuel to glucose for the brain, potentially improving mitochondrial efficiency and compensating for impaired glucose metabolism [56–58]. Another interesting possibility is that substituting ketone bodies for glucose as energy substrates may allow glucose to be redirected toward other functions in which it serves an irreplaceable role [59]. Ketogenic diets have been used clinically since the early 1920s, primarily as a treatment for epilepsy [60, 61]. These diets have been found to normalize neural dynamics in patients with epilepsy by reducing the frequency of transitions among states [62]. In more recent decades, a number of studies have examined the neuroprotective effects of ketogenic diets on age-related cognitive decline and neurodegenerative disease [63]. These studies have demonstrated that ketosis supports increased brain network stability [51] and can compensate for impaired glucose metabolism in individuals with cognitive impairments [64]. The effects observed here are therefore broadly consistent with the possibility that metabolic support during withdrawal promotes adaptive reorganization of large-scale neural dynamics.

The observed associations with BHB also raise the possibility that exogenous ketone supplements could provide a targeted approach for investigating the effects of ketone availability during alcohol withdrawal. Ketone supplements can increase blood BHB levels without requiring the substantial dietary changes needed to achieve nutritional ketosis, thus allowing the effects of ketone availability to be distinguished from other metabolic consequences of a KD. Preclinical studies have shown that exogenous ketone supplements, similar to a KD, can reduce alcohol withdrawal in rodents [65]. In humans, acute ketone supplement intake has been shown to shift brain energy utilization from glucose toward ketones and reduce alcohol craving in individuals with AUD [66], as well as reduce alcohol liking and wanting following alcohol administration in healthy volunteers [67]. Together, these findings support further investigation of whether increasing BHB levels with exogenous ketones is sufficient to influence large-scale brain network dynamics and alcohol cue reactivity, and could inform the development of metabolic interventions for AUD.

Several limitations are relevant to this work. First, given the relatively small sample size, it will be important to verify these results with additional participants in the future. Second, although the randomized dietary intervention provides an experimental manipulation of diet, the relationship between changes in network organization and changes in alcohol wanting was correlational. Thus, the present findings do not establish a causal pathway from metabolic changes through network organization to craving. Third, although changes in serum BHB levels were associated with VE1, the analyses presented here did not establish that the observed changes in TE or the organization of the state transition landscape were mediated by ketone availability, glutamatergic metabolism, or another specific metabolic mechanism. Finally, the network control analysis depends on several modeling assumptions, including the use of a linear, continuous-time approximation of neural dynamics and the definition of brain states according to large-scale functional networks. Although neural dynamics are nonlinear at microscopic scales, recent work comparing linear and nonlinear models of macroscopic brain activity has shown that linear models can accurately approximate resting-state fMRI and intracranial electrophysiological dynamics, providing empirical support for the model used here [36]. Control energy in this framework should therefore be interpreted as a model-derived measure of the cost of state transitions rather than as a direct measure of metabolic energy consumption by the brain. Nevertheless, prior work has demonstrated correspondence between control energy and regional glucose metabolism measured with FDG-PET, suggesting that model-derived control energy may capture aspects of the brain’s underlying metabolic energy demands [21].

In conclusion, these findings provide evidence that dietary metabolic modulation during alcohol withdrawal is associated with changes in the organization of large-scale neural dynamics. The KD intervention was associated with greater temporal reconfiguration of directed information flow and a trajectory toward greater organization of the state-transition landscape, and changes in this organization were associated with reductions in alcohol cue reactivity. These results provide a framework for connecting alterations in brain energy metabolism with the network-level dynamics that support cue-driven behavior. More broadly, they suggest that understanding metabolic contributions to recovery from alcohol dependence may require consideration not only of regional brain activity and neurotransmitter systems, but also of how metabolic state influences the organization and flexibility of communication across the whole brain.

## Data Availability

Data analyzed in this study were obtained from the National Institute on Alcohol Abuse and Alcoholism under a data transfer agreement and are not publicly available from the authors. Requests for access to the underlying data should be directed to the original data custodians and are subject to applicable institutional approvals and data use agreements.

## Acknowledgements

This work was supported by National Institutes of Health (NIH) grant F32AA030475 to J.K.B. N.D.V. acknowledges support from the Intramural Research Program of the NIH, and D.S.B. acknowledges support from the Army Research Office MURI program [W911NF2410228]. The content is solely the responsibility of the authors and does not necessarily represent the official views of any of the funding agencies.

## Citation Diversity Statement

Recent work in several fields of science has identified a bias in citation practices such that papers from women and other minority scholars are under-cited relative to the number of such papers in the field [68–76]. Here we sought to proactively consider choosing references that reflect the diversity of the field in thought, form of contribution, gender, race, ethnicity, and other factors. First, we obtained the predicted gender of the first and last author of each reference by using databases that store the probability of a first name being carried by a woman [72, 77]. By this measure (and excluding self-citations to the first and last authors of our current paper), our references contain 11.16% woman(first)/woman(last), 22.38% man/woman, 21.56% woman/man, and 44.9% man/man. This method is limited in that a) names, pronouns, and social media profiles used to construct the databases may not, in every case, be indicative of gender identity and b) it cannot account for intersex, non-binary, or transgender people. Second, we obtained predicted racial/ethnic category of the first and last author of each reference by databases that store the probability of a first and last name being carried by an author of color [78, 79]. By this measure (and excluding self-citations), our references contain 13.39% author of color (first)/author of color(last), 13.46% white author/author of color, 22.17% author of color/white author, and 50.98% white author/white author. This method is limited in that a) names and Florida Voter Data to make the predictions may not be indicative of racial/ethnic identity, and b) it cannot account for Indigenous and mixed-race authors, or those who may face differential biases due to the ambiguous racialization or ethnicization of their names. We look forward to future work that could help us to better understand how to support equitable practices in science.

