## Supplementary Figure 1 for "Metabolic modulation of brain network dynamics during alcohol withdrawal"

### Supplementary Information for: Metabolic modulation of brain network dynamics during alcohol withdrawal

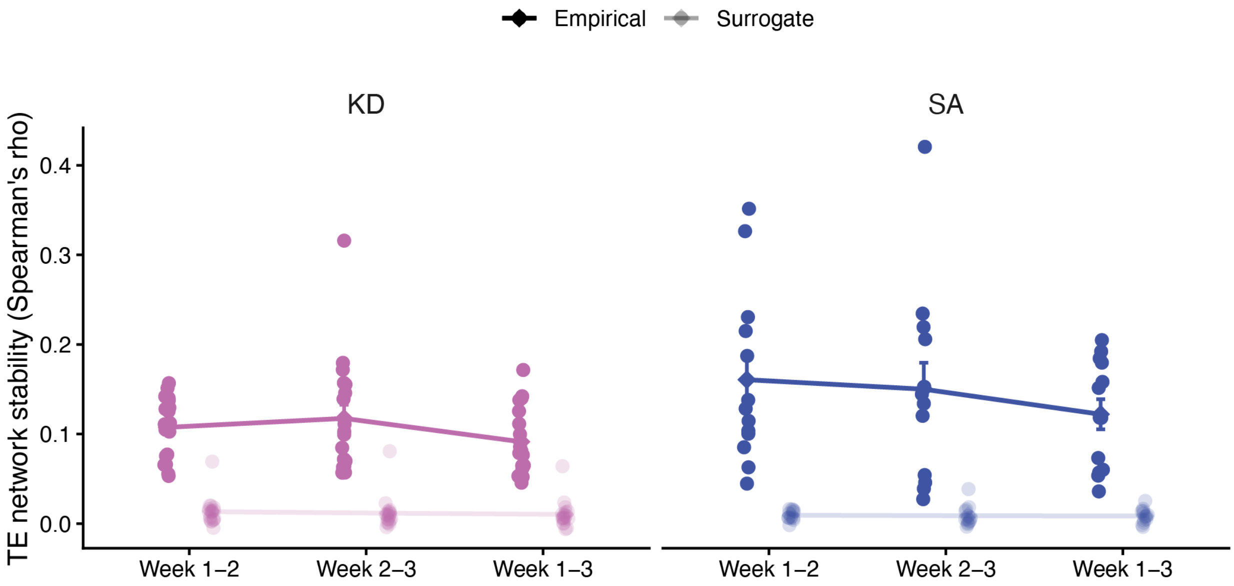


**Supplementary Figure 1: Validation of temporal structure in transfer entropy networks.** Participant-level TE network stability for empirical and circular-shift surrogate networks across weeks 1–2, weeks 2–3, and weeks 1–3 in the ketogenic diet (KD) and standard American diet (SA) groups. For each fMRI run, regional BOLD time series were independently circularly shifted prior to recalculation of transfer entropy (TE). This procedure preserved the temporal structure of each individual regional signal while disrupting temporal alignment between regions. Empirical and surrogate TE matrices were then averaged across available runs within each participant and week, and network stability was quantified as the Spearman correlation of directed edge weights between weeks. Across both treatment groups, empirical networks exhibited substantially greater temporal stability than surrogate networks (main effect of network type, $F(1, 174) = 211.299$, $p < 0.0001$). The magnitude of the empirical-surrogate difference differed between treatment groups (network type $\times$ group interaction, $F(1, 174) = 9.470$, $p = 0.0024$), consistent with greater temporal stability of the empirical TE topology in the SA group. There was no network type $\times$ group $\times$ interval interaction ($F(2, 174) = 0.459$, $p = 0.6327$). Points represent individual participants; summary values represent mean $\pm$ SEM.
